# COVID-19 immunization threshold(s): an analysis

**DOI:** 10.1101/2021.01.02.20248596

**Authors:** Luis Alfredo Bautista Balbás, Mario Gil Conesa, Blanca Bautista Balbás, Ainhoa Alcaide Jiménez, Gil Rodríguez Caravaca

## Abstract

As COVID-19 vaccine research efforts seem to be yielding the first tangible results, the proportion of individuals needed to reap the benefits of herd immunity is a key element from a Public Health programs perspective.

This magnitude, termed the critical immunization threshold (*q*), can be obtained from the classical SIR model equilibrium equation, equaling (1 − 1*/R*_0_*)/ ϵ*, where *R*_0_ is the basic reproduction number and *ϵ* is the vaccine efficacy. When a significant proportion of the population is already immune, this becomes *(n* − 1*/R*_0_*)/ ϵ*, where *n* is the proportion of non-immune individuals. A similar equation can be obtained for short-term immunization thresholds(*q*_*t*_), which are dependent on *R*_*t*_.

*q*s for most countries are between 60-75% of the population. Current *q*_*t*_ for most countries are between 20-40%.

Therefore, the combination of gradual vaccination and other non-pharmaceutical interventions will mark the transition to the herd immunity, providing that the later turns out to be a feasible objective. Nevertheless, immunization through vaccination is a complex issue and many challenges might appear.

## 3 Introduction

Herd immunity is a phenomenon observed when a sufficiently large percentage of a population is immune to an infectious disease, hence reducing the risk of infection of the non-immune individuals. This is related to the fact that a significant proportion of contacts of an infectee will not be susceptible to the disease. When this phenomenon is present, contagions or limited outbreaks or might still take place but large-scale sustained or exponential transmission will not occur.

Herd immunity cutoff (*H*) is classically calculated by *H =* 1 − (1*/R*_0_)^1^ where *R*_0_ is the basic reproduction number, the number of infections each infected case is expected to cause when all the population is susceptible and no special measures are implemented. If the vaccine provides partial protection,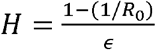 where *ϵ* is the proportional vaccine efficacy. The *R*_0_ of infectious diseases can vary widely between populations due to cultural behaviors, population density and other factors, so local estimates should be used if available; for example in measles it varies between 12-18.^2^

In the context of the COVID-19 ongoing pandemic, herd immunity was proposed as a possible protection mechanism. However, relying on herd immunity has a number of issues due to the lacking evidence about long-term protection^3^. As the results of phase II/III trials of COVID-19 vaccines are made available^4^, the discussion now pertains to the challenges of achieving herd-immunity through vaccination^5^.

In the SIR compartmental models of epidemic diseases, individuals are classified in three compartments: susceptible, infected and recovered; *S, I* and *R* are their respective proportions, (*S + I + R =* 1). As the epidemic unfolds the number of individuals in the *S* compartment decreases, since these individuals are moved to other compartments. We assume there is not any shift from the *R* compartment to the *S* compartment (immunity loss), as this would add a currently unknown rate to the model. In any case, the reproduction number over time can be calculated by *R*_*t*_ *= R*_0_*S*_*t*_ where *S*_*t*_ is the proportion of susceptible individuals at time *t*. If compartmental models are specified with immunity loss (SEIRS model), some periodic oscillations are observed until equilibrium is reached^6^.

This article includes a small theoretical analysis regarding short and long term herd immunity in relation to reproduction numbers, in partially immune populations, and the derived interpretations in the context of COVID-19.

## 4 Materials and methods

The herd immunity cutoff can be obtained via the SIR equilibrium, which is reached when *R*_0_*S*_*e*_ *= 1* (where *S*_*e*_ is the proportion of susceptible individuals at the equilibrium). For a partially-immune population in which vaccination will be introduced, assuming both vaccination and positive antibodies provide lifelong immunity, *S*_*e*_ can be expressed as *S*_*e*_ *= 1* − *qϵ* − *g* where *q* is the *critical immunization threshold* (the proportion of the population that should be vaccinated to reach the equilibrium, where herd immunity starts to act), *g* is the seroprevalence (proportion of already immune individuals) before vaccination and *n* the proportion of non-immune individuals, so that *1 = g+ n*. From this, the *q* would be:

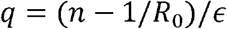

Effective short-term herd immunity can also be obtained as a function of current estimated effective reproductive numbers (*R(t)*) at time *t* in time-since-infection models, which can be calculated by the methods discussed by other authors^7^,^8^,^9^. The instantaneous reproduction number 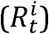 is the number of individuals each individual infected at time *t* is expected to infect, should conditions remain the same. It can be descomposed in: 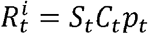, where *C* is the contact rate of infected individuals at time *t* (thus influenced by events restrictions, social distancing, etc.), and *p*_*t*_ is the infection probability of their contacts (thus influenced by wearing masks, increasing ventilation, etc). 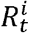, *C*_*t*_ and *p*_*t*_ would be applicable to the individuals that got infected at *t* if conditions remained the same^7^, and more precisely, *C*_*t*_ and *p*_*t*_ would be the average *C* and *p* of the infected population at *t*, both weighted by individual infectivities^9^ at that time point. The case reproduction number 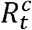 is the number of individuals each infected individual at time *t* actually infects, so it would be 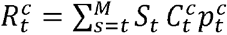, with time-dependent *S* proportion, actual cases contact rates 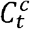 and actual case infection probabilities 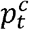 at each time between *t* and, being the number of periods in which an individual is infective. (In more elaborate analysis *N* transmission routes could be used (with their *N* respective contact rates and *N* infection probabilities), so that *C*_*t*_ *\* p*_*t*_ would be a case of matrix multiplication delivering the number of infective contacts).

Let the vaccination be considered at time *t*, with a partially immune population. Equilibrium would be reached when 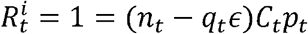, where *n*_*t*_ is the proportion of non-immune individuals at *t* and *q*_*t*_ the proportion of the population that should be vaccinated to reach the equilibrium, should future *C*s and *p*s remain the same. *C*_*t*_*p*_*t*_ can be calculated for any previous time period (*u* period(s) before *t*), and used assuming circumstances will be similar, based on knowledge of cultural habits, contact rates and interactions: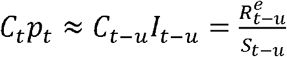. From this:

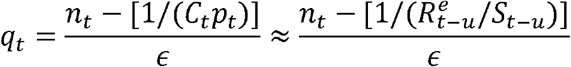

The same can be applied to the *R*_0_ from the SIR model: *R*_0_ *= C*_0_*p*_0_ where *C*_0_ is the average number of contacts during the the infectious period and *p*_0_ the infection/contact probability. *C*_*t*_*p*_*t*_ can be understood as the *R*_0_ we would estimate via a SIR model from a fully susceptible population with contact rates, infectiousness profiles, habits, etc. being the same as in *t*: In this case, *R*_0_ *= C*_0_*p*_0_ *≈1 C*_*t*_*p*_*t*_. The relationship between *q* and *R*_0_ is also similar to that between *q*_*t*_ and 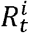, probably because the underlying mechanism is the same.

We have obtained graphics displaying the relationship between *R*_0_ and *q, R*_*t*_ and *q*_*t*_, and *C*_*t*_*R*_*t*_ and 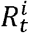 by using several common *E* and *S* values, and calculated the *q* estimates for several countries/situations. Seroprevalence has been assumed by dividing total reported cases (from the COVID-19 CSSE-JHU dataset^10^) by total population (from WHO world population prospects^11^). As *R*_0_ estimates, we used the median (3.32) from the meta-analysis by Alimohamadi et al^12^ and country estimates from Hilton et al^13^.

## 5 Results

The *q* of a population in which 90% individuals are susceptible and the vaccine has 95% efficacy, would be 68.3% if *R*_0_ *= 3*.*32* and 57.9 if *R*_0_ *= 2*.*5*. Figure 1 displays the *q* curves as a function of *R*_0_, *ϵ* and *g*.

**Figure 1:**
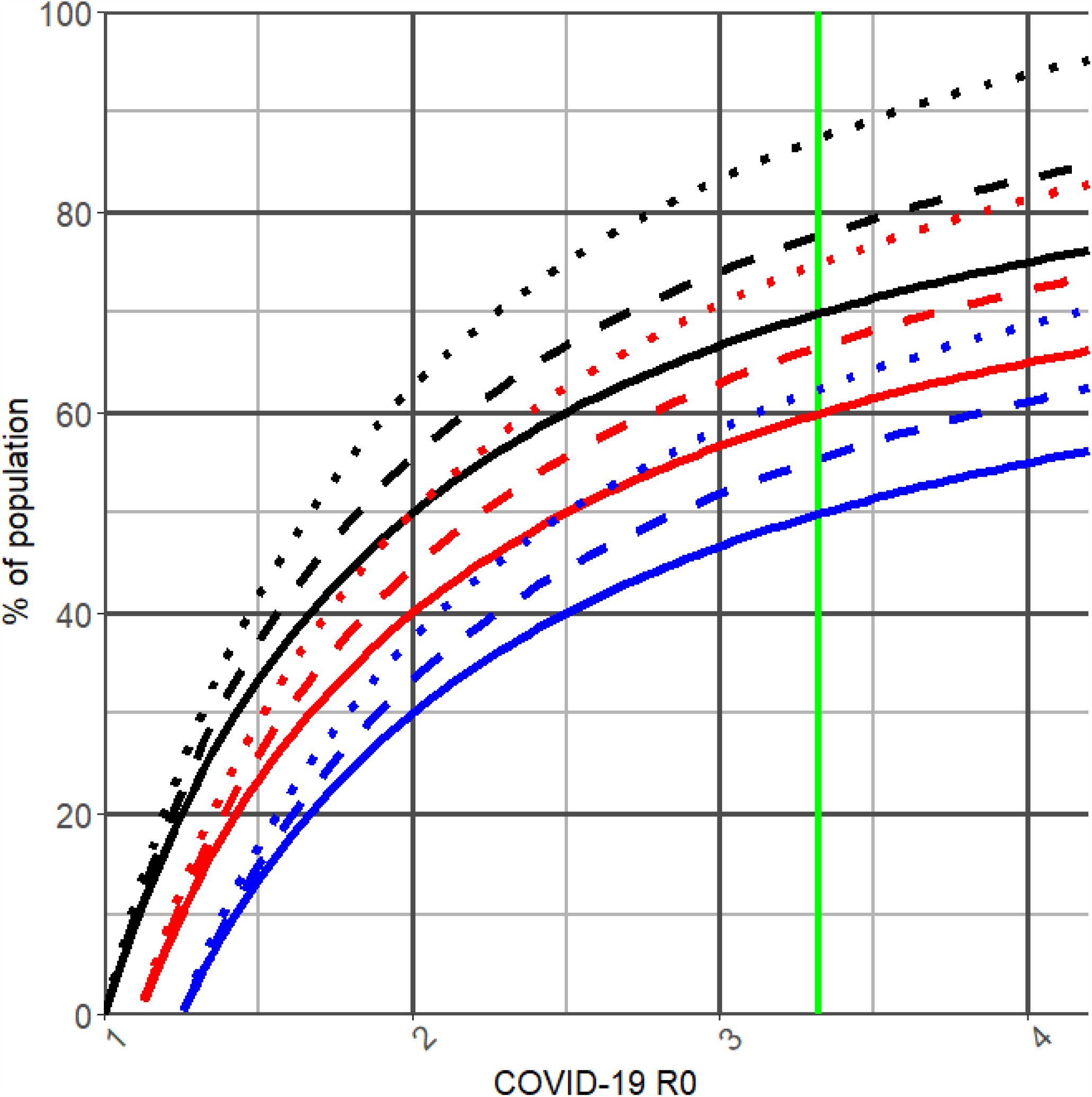
Critical Immunization Thresholds calculated for several situations. Continuous line: 100% vaccine efficacy (e); dashed line: 90% vaccine efficacy, dotted line: 80% vaccine efficacy. Black: 0% population is already immune (g); red: 10% population is immune; blue: 20% population is immune. Vertical green: R0=3.32

The *q*_*t*_ of a population in which *S =* 95% and *E =* 0.95, would be 12.3% if *R*_*t*_ *= 1*.*2* and 29.8 if *R*_*t*_ *=* 1.5. Figure 2 displays the *q*_*t*_ curves as a function of *R*_*t-1*_, *ϵ* and *g*_*t-1*_. Figure 3, shows *R*_*t*_ as a function of *S*_*t*_ and *C*_*t*_*P*_*t*_.

**Figure 2:**
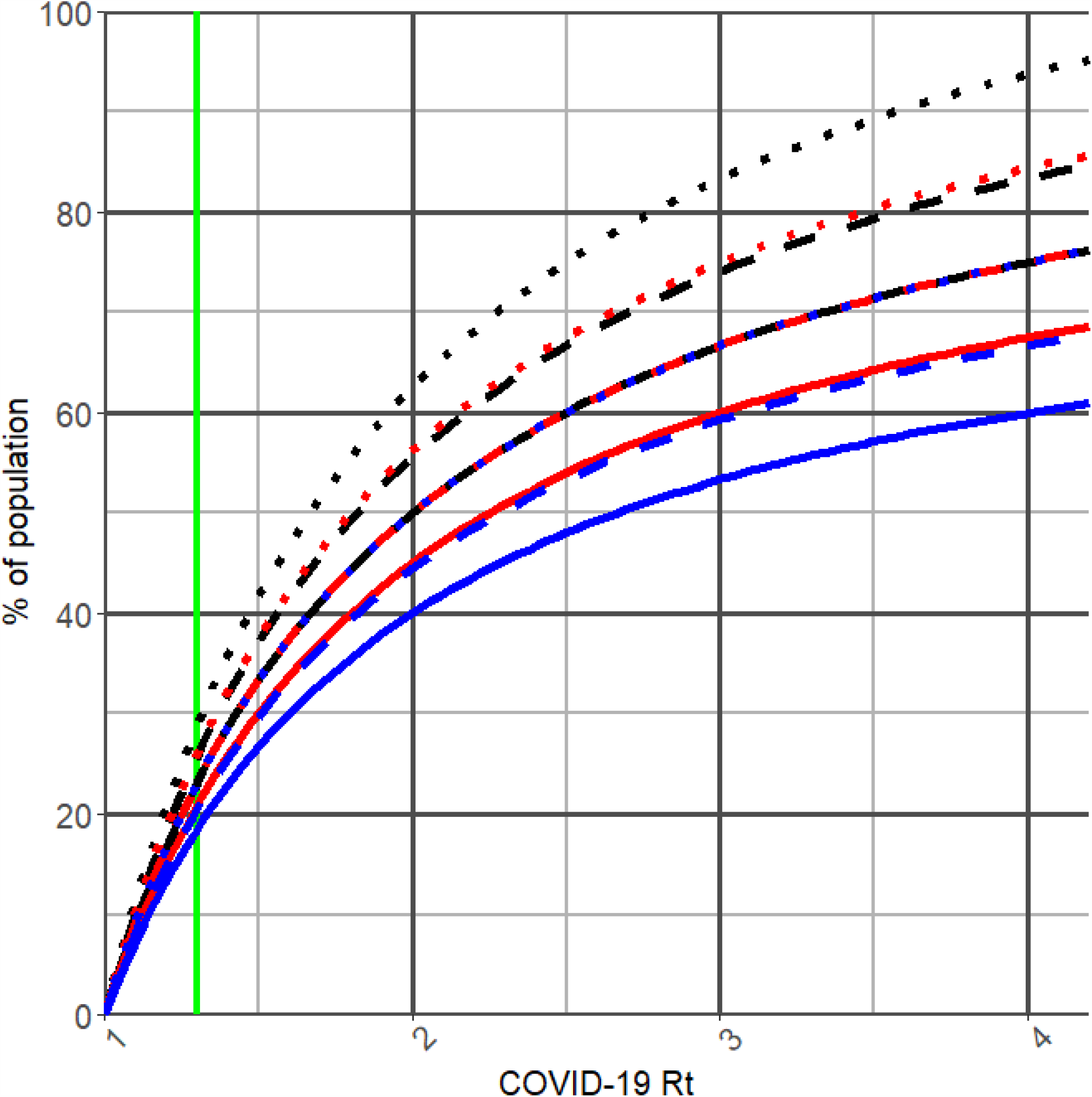
Short-term Critical Immunization Thresholds. Continuous line: 100% vaccine efficacy (e); dashed line: 90% vaccine efficacy, dotted line: 80% vaccine efficacy. Black: 0% population is already immune (g); red: 10% population is immune; blue: 20% population is immune.

**Figure 3:**
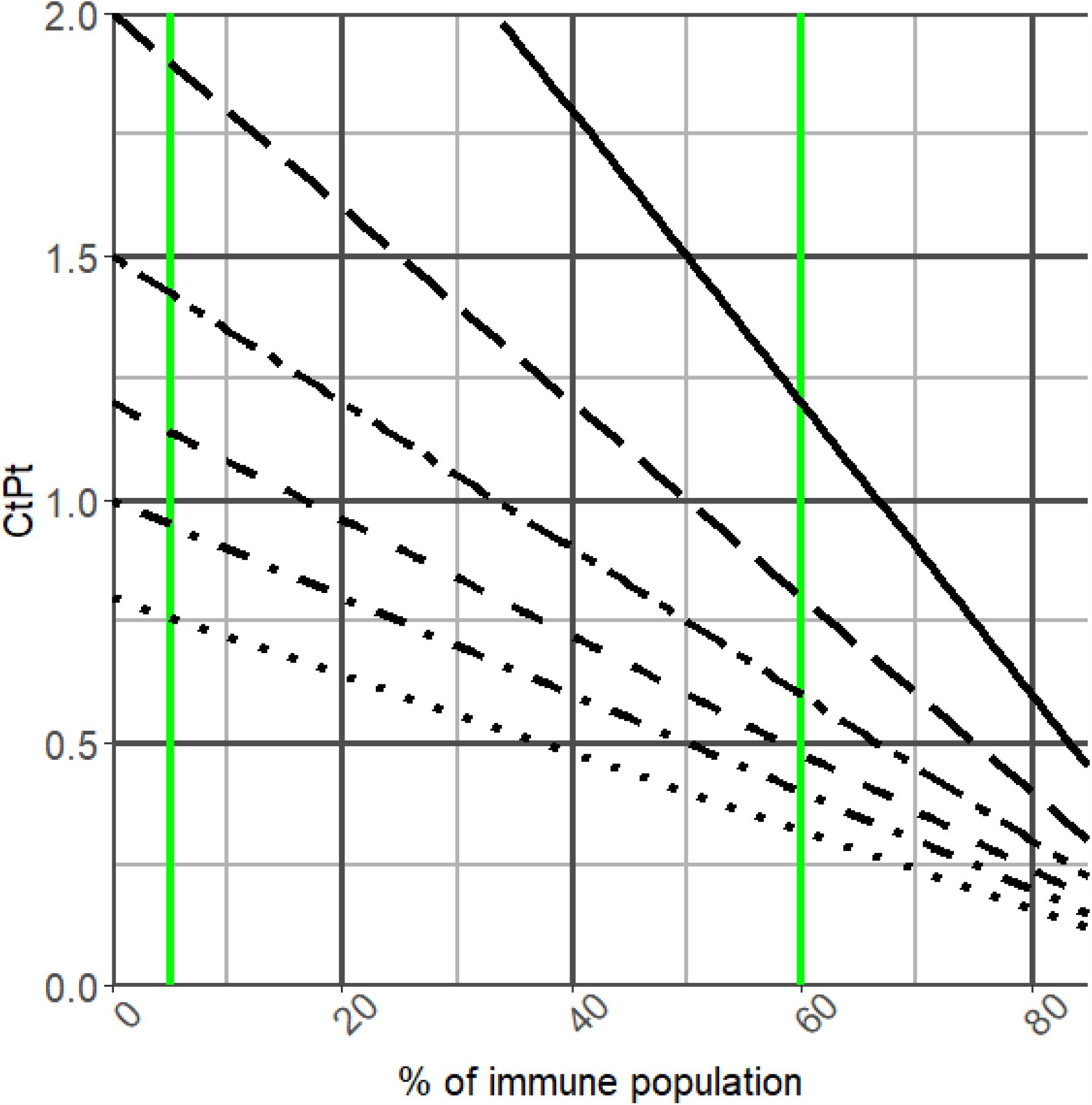
Rt as a function of C_t_P_t_ (reflecting contacts, habits and preventive measures) and susceptible population (S_t_). Dotted line: C_t_P_t_ = 0.8; dotdash: C_t_P_t_ = 1; dashed: C_t_P_t_ = 1.2; two dashes: C_t_P_t_ = 1.5; long dashes: C_t_P_t_ = 2; continuous line: C_t_P_t_ = 3. Vertical lines: 5% seroprevalence, 60% seroprevalence. As the proportion of susceptible population increases, preventive measures can be relaxed while maintaining epidemic control. If transgressions (sudden increases in C_t_P_t_) take place, if a population immunity is small the outbreak will probably be stronger.

Table 1 displays the *q* calculated for several large countries.

**Table 1:** *q*s for several countries, calculated with per-country *R*_0_ (from Hilton et al) or with a global estimate (3.32, from Alimohamadi et al). Number of reported cases as of 01-Jan-2021 is taken as the proportion of immune individuals.

| Country | $R_0$ | Immune proportion | $q$ |
| --- | --- | --- | --- |
| United States of America | 2.22 | 6.1 | 48.85 |
| United States of America | 3.32 | 6.1 | 63.78 |
| India | 2.78 | 0.7 | 63.33 |
| India | 3.32 | 0.7 | 69.18 |
| Brazil | 2.79 | 3.6 | 60.56 |
| Brazil | 3.32 | 3.6 | 66.28 |
| Russian Federation | 2.18 | 2.2 | 51.93 |
| Russian Federation | 3.32 | 2.2 | 67.68 |
| Mexico | 2.65 | 1.1 | 61.16 |
| Mexico | 3.32 | 1.1 | 68.78 |
| Sweden | 2.14 | 4.3 | 48.97 |
| Sweden | 3.32 | 4.3 | 65.58 |
| United Kingdom | 1.67 | 3.7 | 36.42 |
| United Kingdom | 3.32 | 3.7 | 66.18 |
| Spain | 2.00 | 4.1 | 45.90 |
| Spain | 3.32 | 4.1 | 65.78 |
| France | 2.01 | 4.0 | 46.25 |
| France | 3.32 | 4.0 | 65.88 |
| Ukraine | 2.15 | 2.5 | 50.99 |
| Ukraine | 3.32 | 2.5 | 67.38 |

## 6 Discussion

The *critical immunization threshold* depends on *R* number, percentage of susceptibles and vaccine efficacy. Many estimates are available for COVID-19 *R*_0_; each model having its own assumptions. These were obtained from early pandemic periods with partial implementation of protective measures, therefore maybe underestimating *R*_0_.

The proportion of immune individuals varies between countries. We used the number of reported cases, a conservative approach, given the under-reporting of cases. Reported cases range between 0-5% population in most countries. The initial seroprevalence study in Spain (April-May 2020) show similar ranges (5%)^14^. In 3609 adult volunteers from Liguria and Lombardia, a rate of 11% (95%CI: 10.0-12.1%) was observed^15^. In Kenyan blood-donors, up to 8% seroprevalence was observed in some urban counties^16^.

Actual future vaccine efficacy can only be subject to conjecture at present. The BNT162b2 mRNA Covid-19 Vaccine (by Pfizer®) has shown a 95% (95%CI: 90.3-97.6) protection against COVID-19 infection^4^. With the ChAdOx1 nCoV-19 vaccine (AZD1222), efficacy was 62.1 (95%CI 41.0–75.7) for the two-standard-doses regime, and 90.0% (67.4–97.0) for those who initially received a low dose instead^17^.

By using this data, COVID-19 *q* would be 60-70%. If we calculate *q*_*t*_ estimates for *R*_*t*_s under 1.5, which is the case for developed and developing countries after the first wave of the pandemic;^18^ these would lie around 20-40%.

An important consequence of these graphs is that, at equilibrium, initially small variations of *R* are compensated by large proportions of vaccinated individuals, suggesting the benefits of vaccines might not be apparent at the initial periods.

This calculation has several limitations. We have used epidemiological models that do not take into account population heterogeneity and the non-random habits of particular mixing groups, an important limitation of herd immunity estimations^19^. As vaccination and contacts will not be random, this discussion is and approximation. Additionally, *R*_0_ is always an estimate, and considerable differences can be observed between studies, rendering *q* calculation uncertain. Population growth, migrations and deceases have not been included. (If pre-pandemic population is used, newborns are susceptible, and infected-and-deceased should be excluded from the immunes).

Finally, vaccination and immunity is a complex process: Virus mutations and strains, vaccine hesitancy, efficacy in high-risk groups, protection duration, mucosal immunity, etc..^5^ Evidence is scarce regarding the duration of COVID-19 immunity^3^, and different kinds of immunity against other coronavirus might wane over years^20^. Several rare cases of reinfections have been reported^21^, but still orders of magnitude smaller than the number of new infections; and booster vaccine doses might be required.

As advantages, this study can help better understand the road lying ahead. In the short term, the *R*_*t*_ will be kept lower than 2 in many areas due to contact restrictions, remote working, etc. Despite the public and private efforts, there is uncertainty regarding the number of vaccine courses available by the end of 2021 for many countries^22^; in this context effective epidemic control can be achieved through a combination of preventive measures and gradual population vaccination^23^. On the other hand, vaccine hesitancy has been observed in different countries (USA^24^, Turkey^25^,…), with undecided (∼30%) or refusing individuals (∼10%), and concern exists regarding their management. These percentages are not contradictory with reaching high percentages of immunity, and gradual vaccination campaigns can allow these individuals to make an informed decision. It should be noted vaccination strategies include different objectives: maintaining essential services (targeting healthcare, police and other workers), reducing severe cases (targeting comorbidities or the elderly), or stopping the virus transmission (targeting other individuals)^26^. As a lesser-priority group, asymptomatic COVID-19 cases with weaker immune response might benefit from receiving a booster dose^27^.

Even if the *q* is not reached, reducing the susceptible population has advantages regarding smoother and less sudden epidemic outbreaks. Nevertheless, achieving *q* value through vaccination doesn’t guarantee any lack of new contagions: if coverage varies between groups in a population, those with insufficient immunization can exhibit sustained transmission^1^.

To conclude, we have illustrated the effect of population immunity in epidemic control is quantitative, and interdependent with contact rates and their determinants.

## Data Availability

Full R code will be made available upon manuscript final publication

